# Preparedness in practice: An outbreak science approach to studying public health emergency response

**DOI:** 10.1101/2023.06.24.23291861

**Authors:** Mackenzie Moore, Hailey Robertson, David Rosado, Ellie Graeden, Colin J. Carlson, Rebecca Katz

## Abstract

Outbreak response, as a technical and specialized field of practice, is struggling to keep pace with the evolving dynamics of modern public health emergencies. Extensive scholarship across disciplines and epidemics has highlighted the importance of early action, the costs associated with delayed mobilization, the necessity of effective preparedness plans for complex crises, and the growing need for response to operate in spite of both uncertain information and social disruptions. Here, we present and analyze a new dataset of 235 different multisectoral activities that comprise outbreak preparedness and response. We explore the conditions under which these activities are applicable, including different phases of response, different operating circumstances, and different disease etiologies, and find that the core activities required for outbreak response largely apply across etiology and scale, but are more substantial during the early phases of response. To validate this framework with real-world examples, we then examine 279 reports from the WHO Disease Outbreak News (DON), a narrative record of outbreak history through time, and examine which of our activities are reported or implied in these narratives. We find that the core components of response are applicable across the vast majority of biological events, especially as they relate to basic epidemiology, infection prevention, and governance, and that many different kinds of real-world outbreaks require the same core set of responses. These findings point to a nearly-universal set of outbreak response activities that could be directly incorporated into national and international response plans, significantly reducing the risk and impact of infectious disease outbreaks.

## INTRODUCTION

Outbreak response requires rapid, cross-sector coordination under high-consequence, complex conditions, all while operating with limited information. Early and effective response can not only reduce the impact of outbreaks, but under the right circumstances, can entirely prevent them from progressing into epidemics or pandemics. However, during the Covid-19 pandemic especially, decision-makers conspicuously struggled to prioritize and act (Singh et al. 2021). Despite a plethora of preparedness plans and frameworks that had been developed following earlier outbreaks (e.g., the SARS outbreak, the H1N1 influenza pandemic, multiple epidemics of Ebola virus, and the Zika epidemic in the Americas), these plans often failed to translate into real-world policies or interventions, with an undeniable cost to human health and well-being (Haldane et al. 2021). It will take decades to sift through these failures, in large part due to the difficulty of defining the counterfactual for what “should” or “could” have happened at each stage of decision-making and mobilization. Even more challenging is to compare *across* outbreaks and outbreak types, in order to identify failures and weaknesses that are shared between events—and to distinguish individual, momentary failures from structural weaknesses in the global health security architecture.

Understanding the steps taken to identify, intervene in, and mitigate the impacts of disease outbreaks is one of the core components of outbreak science (Rivers et al. 2019), but so far, it has received less attention than other quantitative aspects of outbreak analytics. To address this gap, in this study, we analyze 235 outbreak-related activities that span sectors and stages of outbreak response, and develop a taxonomy for how to categorize their scope, intentions, and relevance to different circumstances. We take a “whole-of-society” approach, aiming to capture as broad a set of sectors and activity types as possible, well beyond the narrow powers of public health agencies and the responsibilities of boots-on-the-ground epidemiologists. We similarly take as broad a “temporal” view as possible, starting with the systems that first detect outbreaks, and continuing into the outbreak recovery phase, sometimes lasting years after transmission ends. Using this activity dataset, we examine applicability to different hypothetical scenarios under which a response might need to be conducted. To validate our framework—and test its real-world relevance—we next examine which of these activities were present in a sample of 279 real-world reports from the WHO Disease Outbreak News, spanning 1996 to 2019 (Carlson et al. 2023). Using this comparison, we explore how these kinds of outbreak histories capture the basic components of response, and whether the most common real-world response activities differ from those identified by our exercise. We conclude by briefly discussing the implications for future efforts to standardize outbreak preparedness plans.

## METHODS

### The activity dataset

The Georgetown Outbreak Activity Library (GOAL; outbreaklibrary.org) is an established resource for public and professional reference on outbreak response (Katz and Graeden 2020). The contents of GOAL were developed over several years, based on collaborative workshops; guidance from partners at the World Health Organization (WHO), the Emergency Response Frameworks from the WHO, the United Nations Office for the Coordination of Humanitarian Affairs, the United Nations Children’s Fund, and international non-governmental organizations; peer-reviewed and technical literature; and epidemiology textbooks. Based on these sources, the GOAL website (and underlying dataset) captures a total of 229 outbreak response activities, which fall into six broad categories: epidemiology & lab analysis; healthcare and infection, prevention, and control (IPC); governance and coordination; humanitarian assistance; safety and security; and logistics and support. On the website, each activity has a brief description, with links to various case studies, policies, and other documents.

### Categorizing outbreak response activities

Here, we use the GOAL project as a reference dataset on the components of outbreak response. Using a combination of the previously-listed literature sources and our expert opinion, we developed a dataset of the contexts in which each of the 229 outbreak response activities could plausibly be applicable, with a focus on three areas: geographic scale, outbreak phase, and disease etiology. While individual activities’ inclusion or granularity in GOAL can be somewhat subjective, the comparative framework we apply is the more salient contribution of this effort.

To describe the geographic scale at which an outbreak activity was applicable, we used a simple grouping of *subnational, national*, and *international*, with an eye towards international organizations’ involvement as an often-neglected enabler of response activities.

To describe the timeline over which activities take place during an outbreak, we developed a simple categorical taxonomy of *outbreak phases*. Based on previous literature about the idea of the “epidemiological curve” (Polonsky et al. 2019) and the stages of disease emergence (Lloyd-Smith et al. 2009; Wolfe, Dunavan, and Diamond 2007), we identified five outbreak phases: *surveillance & preparedness* (baseline operations before an outbreak begins), *detection* (when an outbreak is first identified and investigated), *early response* (usually primarily focused on containment), *intervention* (full-scale, real-time response during an ongoing or worsening event), and *post-intervention & recovery* (response wind-down, after-action reports, and efforts to rebuild and mitigate social and economic impacts). These often collectively form a cycle, as post-outbreak recovery ideally leads into efforts to improve preparedness and reduce the risk of future outbreaks (Phelan and Carlson 2022)

Finally, we developed a *etiological taxonomy* to categorize activities by the type(s) of disease origin and transmission that are targeted by outbreak response. This taxonomy reflects both the true underlying etiology of an outbreak, and the level of biological and epidemiological information available to responders. For infectious diseases, emerging diseases tend to follow a progression from animal-only to limited- and eventually ongoing human-to-human transmission, sometimes with the help of arthropod vectors (Lloyd-Smith et al. 2009; Wolfe, Dunavan, and Diamond 2007; Kreuder Johnson et al. 2015). Other outbreaks have entirely non-communicable etiology (e.g., melamine contamination, radiation exposure), or are caused by an agent that is never identified. Occasionally, new transmission modes can appear over time, such as sexual transmission of familiar vector-borne pathogens (e.g., Zika virus) and zoonoses (e.g., mpox) (Foy et al. 2011; Laurenson-Schafer et al. 2023). The scientific task of outbreak response generally involves pinpointing these dynamics as the relevant data become available, but often lags behind ground truth: for example, it may take days or weeks to confirm that human-to-human transmission is occurring after a novel zoonotic disease emerges, even if an epidemic is steadily growing (Singh et al. 2021). Our etiological taxonomy was used to describe which situational reality outbreak responders would *believe* they are responding to, based on available information, and was divided into ten categories (Table 1).

**Table 1.** A situational taxonomy for outbreaks defined by available information on disease etiology.

| Event type | Description |
| --- | --- |
| Localized syndromic event | A cluster of cases with similar symptoms and an unknown causative agent. Once more information is available, it is expected but not guaranteed that the event will be reclassified (e.g., acute febrile illness or nodding disease) |
| Localized exposure | Early indications of a cluster of individuals seemingly linked to a common exposure to a disease-causing agent (e.g., radiation poisoning) |
| Zoonotic spillover without known onward transmission | A documented event where a pathogen jumps species from animal to human. The human case, however, does not seem to be able to transmit the pathogen to other humans (e.g., Hendra virus) |
| Zoonotic spillover with human-to-human transmission | A documented event where a pathogen jumps species from an animal to a human, and then there is sustained human to human transmission. It may also be possible for the pathogen to be reintroduced multiple times from animals to humans (e.g., Ebola virus) |
| Vector-borne transmission without known onward transmission | A pathogen transmitted from animals to humans by an arthropod vector, but humans are a dead-end host (e.g., West Nile virus) |
| Vector-borne transmission with onward transmission | A pathogen transmitted primarily from human to human by an arthropod vector (e.g. dengue fever) |
| Vector-borne transmission with human-to-human direct transmission | A pathogen transmitted primarily from human to human by an arthropod vector, but transmission can also be from human to human (eg., pneumonic plague; sexual transmission of Zika virus) |
| Human pathogen with environmental transmission | A pathogen is transmitted through a non-respiratory environmental exposure, including fecal-oral transmission (e.g., *Cryptosporidium*) |
| Human pathogen with respiratory transmission | A pathogen that is transmitted human to human through a respiratory route – either airborne or through droplets (e.g., measles) |
| Human pathogen with transmission via bodily fluids | A pathogen spread between humans through bodily fluids (e.g., HIV) |

### An analysis of the Disease Outbreak News

To validate our analysis with real-world data on outbreak response, we applied this taxonomy for outbreak response analysis to the World Health Organization (WHO)’s Disease Outbreak News (DON). These documents serve as the WHO’s real–time record of notable outbreaks, and is one of the longest-running public sources of data on infectious disease response. Although these reports are generally biased towards larger and especially multi-national outbreaks (i.e., public health emergencies with significant WHO involvement), and are further limited by inconsistencies in both national reporting and WHO decisionmaking about information sharing, the narratives in the DON nevertheless provide a relatively unique set of textual data about outbreak responses and contexts over space and time. Previously, we compiled a systematic index of 2,789 DON reports between January 1996 and December 2019 (i.e., ending prior to the Covid-19 pandemic), with standardized metadata on disease and locality, but limited information on interventions (Carlson et al. 2023).

We randomly selected a ∼10% sample of these data, and – after examining which reports were still accessible – were left with 279 unique entries in the Disease Outbreak News. From the report texts, we identified any activities in our framework that were explicitly recorded or implied (e.g., “Count cases” is implicit in text reporting case totals). Two researchers coded each report, and an additional three researchers reconciled all coding disagreements. From this final sample, we identified six new activities that were consistently or significantly reported enough to merit back-inclusion in the activity taxonomy, which has since been updated to include them. These activities included: “Carry out screening/testing activities”; “Collect case patient information”; “Implement vector control measures”; “Initiate immunization campaign”; “Provide medical countermeasures (MCMs)”; and “Provide technical assistance.” This brought the total number of activities from 229 to 235, and for each of the new activities, we again coded geographic scope, outbreak phase, and relevant etiologies.

### Limitations

The list of activities reflects, in some cases, bias from the research team performing the analysis. Specifically, activities related to governance are represented more heavily than others due to the policy interests and expertise of the authors; however, the heavy representation also reflects the highly detailed and specific nature of governance activities in this context, and highlights the relative importance of including policy and governance expertise in outbreak response management efforts.

Our study is also limited by the characteristics of the WHO Disease Outbreak News as a textual record. These reports typically include a bare bones accounting of outbreak progression, focused on the early stages of outbreak identification and risk assessment. From these sources alone, we are unable to differentiate between activities that failed to be implemented and those that were simply not recorded. As highlighted by recent efforts to investigate failures during the Covid-19 response (Singh et al. 2021), the task of describing and chronicling even a single outbreak response is a massive one—especially when including the full span from preparedness to recovery, and when every relevant sector is included. This limitation highlights a need for better retrospective and real-time documentation of outbreak histories, and our goal here is to present a semi-formal framework that can serve as a foundation for that future work.

## Data and code availability

All data and code to reproduce our analysis are available on Github (github.com/cghss/epiintel).

## RESULTS

### Characteristics of outbreak activities

The outbreak activity dataset lists 235 distinct components of outbreak detection, response, and recovery. These activities span the fundamental (e.g., “Count cases”) to the highly applied (“Promote safe burial practices”), and span geographic scales from location specific (“Identify and train local spokespersons”) to global in nature (“Consider request for investigation into potential breach of the Biological Weapons Convention”). However, we found that the majority of activities may be implemented or coordinated across multiple (n = 224 of 235, or ∼95%) or all geographic scales (n = 178, or ∼76%; Subnational: n = 218; National: n = 231; International: n = 188).

Many activities reflected the unique problems faced by public health institutions, especially during an infectious disease epidemic (“Determine mode of transmission”; “Perform contact tracing activities”). However, many activities were more fundamental to emergency management, and also apply to other types of events, such as natural disasters or terrorism (“Activate emergency operations center”; “Engage the media”). When grouped by outbreak phase, the majority of activities were needed in the early stages of emergency response (Surveillance & preparedness: n = 96; Detection: n = 113; Early response: n = 186; Intervention: n = 176; Post-intervention & recovery: n = 119). However, the majority of activities were represented in more than a single stage (n = 201, or ∼86%), with a small percentage applicable to every stage of outbreak preparedness, response, and recovery (n = 43, or ∼18%).

Activities fell into six broad categories (Governance & coordination: n = 90, Logistics & support: n = 74; Epidemiology & lab analysis: n = 64; Healthcare & IPC: n = 56; Humanitarian assistance: n = 36; Safety & security: n = 30); roughly 41% of activities fall into two or three of those categories (n = 96), with the most common co-occurring together in “Healthcare & IPC” and “Logistics & support” (n = 23). Activity categories shift significantly through outbreak phases (χ^2^ = 40.47, df = 20, p = 0.0044; Figure 1); while most activities occur during early response or acute interventions, epidemiological and lab activities are more common in the surveillance and detection phases, while humanitarian response tends to occur during and after the acute outbreak period. By comparison, we found no evidence that the geographic scale of outbreak activities changed through outbreak phases (χ^2^ = 0.46154, df = 8, p = 0.999) or that different categories of activities were clustered at particular geographic scales (χ^2^ = 2.1482, df = 10, p = 0.995), although notably, a large number of activities relating to governance & coordination (n = 88) and logistics & support (n = 74) can be organized at the national scale.

**Figure 1.**
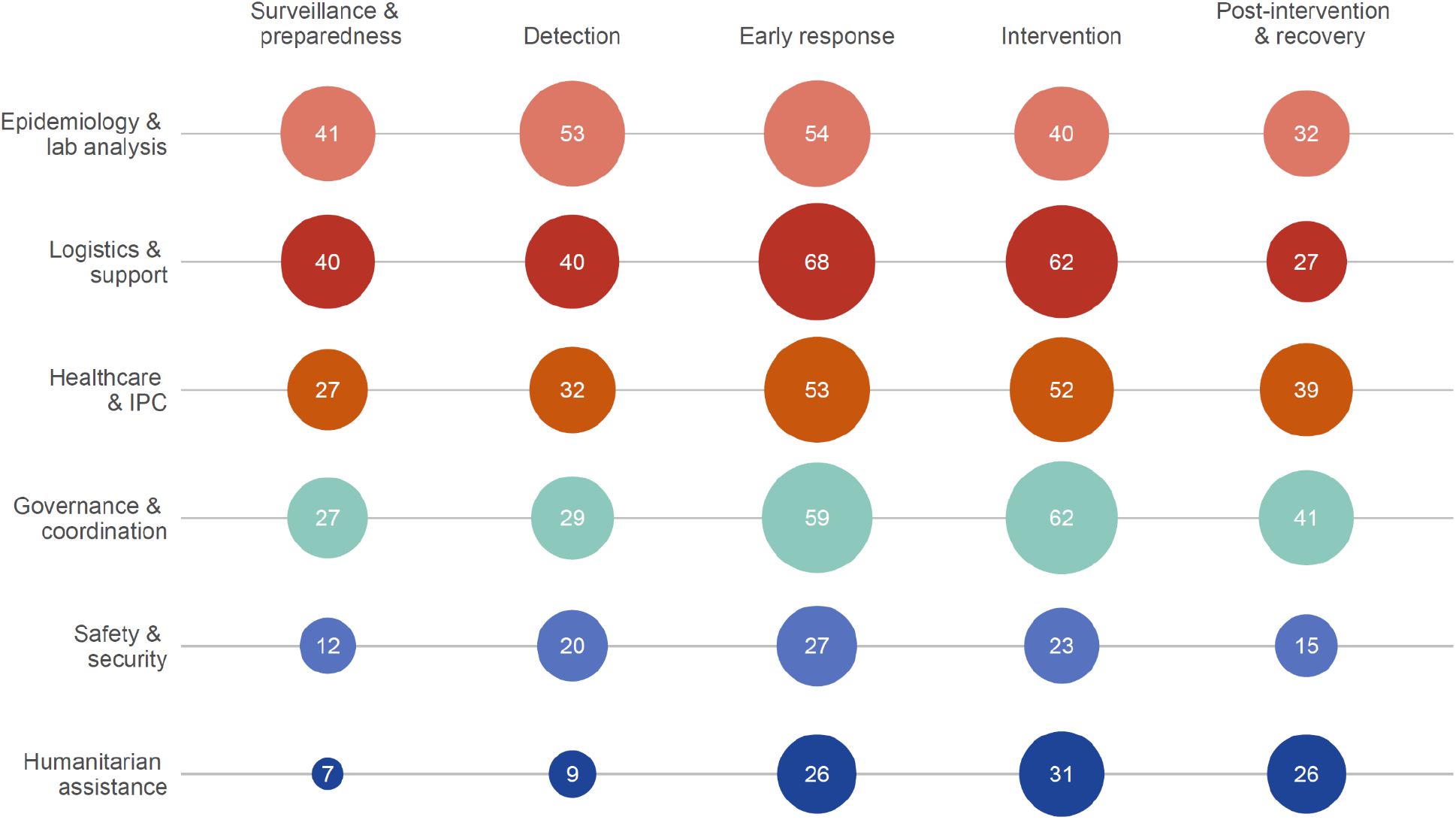
The number of applicable activities (white numbers)in each category shifts through outbreak phases: epidemiological analyses are most applicable in the earliest stages, while response and recovery eventually shift into humanitarian and governance work.

### Comparing activities to hypothetical outbreak scenarios

We found that almost every outbreak activity (n = 233 of 235, or ∼99%) was applicable for every type of biological event, including of a natural, accidental, deliberate, or unknown origin. Only two events were specific to the possibility or presumption of a deliberate event (“Consider request for investigation into potential breach of the Biological Weapons Convention”; “Ensure chain of custody of samples”); even in a deliberate biological event, the fundamental steps required for outbreak detection and containment are mostly unchanged.

Next, we aligned ten basic types of situations (presumed etiologies) with each of the 235 activities and examined whether that activity would be applicable to that type of outbreak. We found that the majority of activities (n = 199, or ∼85%) were applicable in all types of situations, and that every type of outbreak could reasonably require the majority of activities (ranging from dead-end vector borne transmission: n = 209; to zoonotic outbreaks with human-to-human transmission: n = 230). Of the 36 outbreak activities that were not universally applicable across outbreak types, most were applicable to the response or post-response period (n = 35), whereas fewer were relevant to the surveillance and detection phases (n = 21). Specialized activities were disproportionately related to infection prevention or healthcare (n = 20), leaning heavily towards infection prevention measures that involved animal hosts (e.g., “Dispose of animal carcasses, litter, and animal products”; “Vaccinate domestic animal species”), arthropod vectors (e.g., “Implement vector control measures”), and food-borne exposure (“Modify food production or preparation process”; “Close food premises or prohibit the sale or use of foods”), highlighting the value of One Health expertise and the need to engage experts from relevant institutions (e.g., the World Organization for Animal Health).

### Comparing activities to real-world outbreak scenarios

To evaluate the applicability of the activity framework to real-world outbreaks, we cross-referenced our list of outbreak activities against a random sample of 279 outbreak reports taken from the WHO Disease Outbreak News. Although 36 reports included no data on response activities, the majority (n = 243, or ∼87%) did, and we found that more than half (n = 138 of 235, or ∼59%) of our activity list was represented in at least one report. Activities relating to epidemiology (n = 51 of 64, or ∼80%) and infection prevention or treatment (n = 36 of 56, or ∼64%) were most frequently represented, while those related to logistics and support were surprisingly described in less than half of the reports (n = 32 of 70, or ∼43%), reflecting the focus of both the WHO and the Disease Outbreak News itself. Many of the most basic activities were well-represented across reports (Table 2), especially if they related to basic shoe-leather epidemiology (e.g., “Count cases”; “Perform contact tracing activities”). These core activities were generally shared across all types of outbreaks, with a high degree of similarity across epidemics with very different transmission modes, severity, and scales (Figure 2).

**Table 2.** The top 10 activities mapped onto our sample of WHO Disease Outbreak News.

| Activity | Activity categories | Number of DON reports |
| --- | --- | --- |
| 1. Count cases | Epidemiology & lab analysis; Safety & security | 193 |
| 2. Count deaths | Healthcare & IPC | 175 |
| 3. Confirm diagnoses | Epidemiology & lab analysis; Healthcare & IPC; Safety & security | 162 |
| 4. Identify agent based on laboratory testing | Epidemiology & lab analysis; Safety & security | 158 |
| 5. Initiate epidemiological investigation | Epidemiology & lab analysis; Governance & coordination | 112 |
| 6. Provide clinical care | Healthcare & IPC | 88 |
| 7. Coordinate response with international partners | Governance & coordination | 82 |
| 8. Perform contact tracing activities | Epidemiology & lab analysis | 77 |
| 9. WHO/WOAH notification | Governance & coordination | 73 |
| 10. Conduct disease surveillance | Epidemiology & lab analysis | 71 |

**Figure 2.**
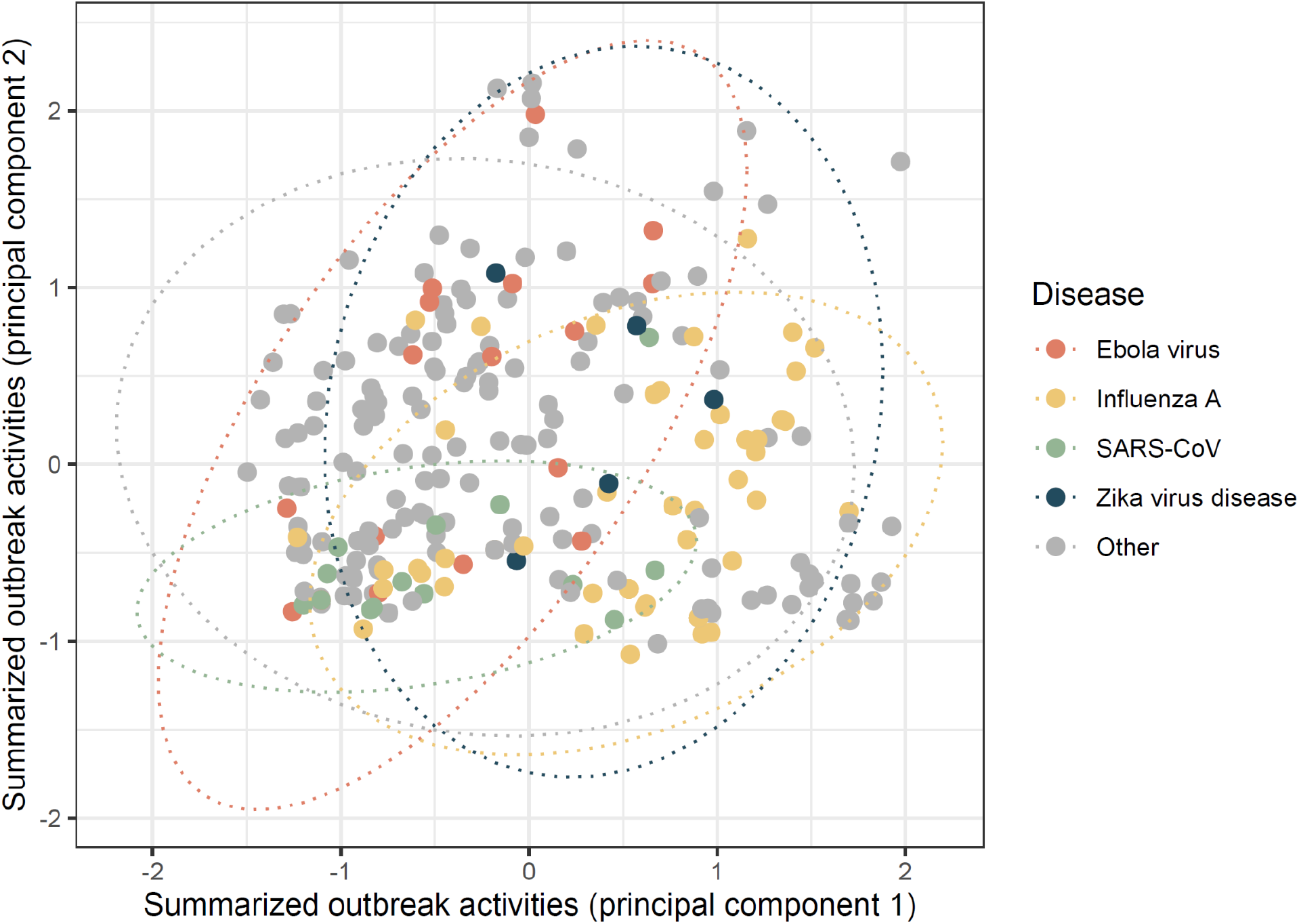
An example visualization of the 243 outbreaks with reported activities, arranged based on a principal components analysis of activities’ binary inclusion, and grouped by specific pathogens of interest. Outbreaks show some segregation in activity space, but with a high degree of overlap: response to most kinds of disease outbreaks relies on the same core set of (at least, reported) activities. (Ellipses show normal distribution estimates of data density by group.)

However, several foundational activities are under-represented. In some cases, these examples are activities that are rare by nature (e.g., “Issue emergency declaration”: only n = 1 report). In others, they fall after WHO stopped reporting on outbreak updates (e.g., “Declare that the outbreak is over”: n = 1, or are steps upstream of more visible activities (e.g., “Determine vaccination policies”: n = 0; and “Identify potential points of dispensing for vaccines and medical countermeasures”: n = 0; versus “Procure and transport medical countermeasures”: n = 22; and “Provide medical countermeasures”: n = 17). Notably, many of the activities that are not indicated by any reports analyzed were those with a humanitarian focus (“Unify separated families”; “Provide food aid”) or targeting human resources (“Train security and law enforcement personnel”; “Support occupational health and safety of responders”). Many of these activities fall at least partially outside the purview of the World Health Organization (which curates and publishes the Disease Outbreak News), and draw in a much broader set of stakeholders that includes the United Nations (UN) Children’s Fund, the UN World Food Programme, the UN Office for the Coordination of Human Affairs, the U.S. Agency for International Development, the World Bank, and International Monetary Fund.

Over several decades, the activities described in the Disease Outbreak News have changed substantially, hinting at the evolving nature of both outbreak response and the textual record itself. Over time, reports have tended to describe a greater number of response activities (Figure 3). This likely reflects a mix of increasing detail in the Disease Outbreak News reports themselves, and an increasingly complex set of information that needs to be shared with countries, media, and the public. In some cases, increasing mention of individual activities speak to major events in the history of global health: for example, after the adoption of the revised International Health Regulations in 2005, more reports begin to describe formal notifications submitted to WHO or WOAH (although the latter category is the subject of a different set of international agreements). However, in other cases — including, notably, the rise of genomic epidemiology — important activities remain under-represented through time, even as they have played an increasingly important role in surveillance and response.

**Figure 3.**
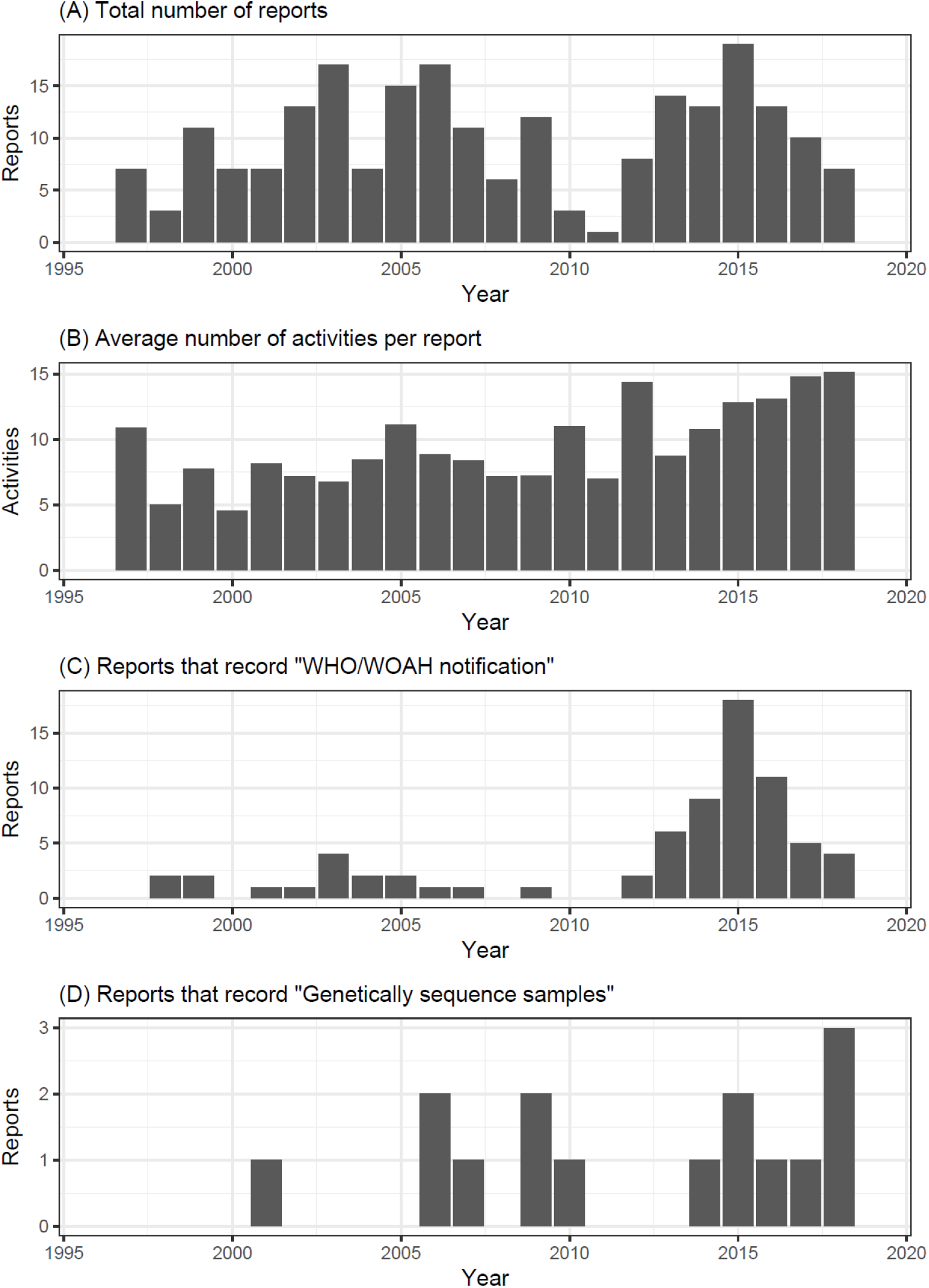
Total number of reports, average number of activities identified based on each report, and two selected activities, in the random sample of WHO Disease Outbreak News that we analyzed.

## DISCUSSION

Here, we analyzed over 200 activities required for outbreak response, and explored a representative sample of real-world outbreaks to validate our framework. We found that while hundreds of activities are required for a whole-of-society approach to outbreak response, the core components of a successful response are shared across different diseases and real-world circumstances. Those minimum core activities generally relate to shoe-leather epidemiology, infection prevention, and resource mobilization, and are enabled by the actions of international organizations but are mostly implemented at the country or local level. In the course of a single outbreak, the required activities may shift from baseline disease surveillance to long-term humanitarian aid; but the vast majority of coordination and mobilization is required in the earliest stages of emergency response.

Taken together, our findings point to the idea that an identifiable set of minimum core capacities exist for outbreak response. Most of the core activities can be initiated in the earliest days of an outbreak: they are generally focused on life-saving measures or information gathering, and are fundamentally local, often with minimal policy barriers to implementation. Because that core response toolkit is mostly shared across different diseases and circumstances, we argue that decision-makers can begin to prioritize, mobilize, and finance a massive number of basic steps on day 1 — even in the absence of more detailed information on the etiology, transmissibility, or severity of a pathogen. In doing so, we argue that public health institutions might better overcome uncertainty and mobilize the rapid response required to keep outbreaks from becoming epidemics or pandemics. That idea is supported by a decade of work focused on timeliness of detection, information sharing, and early response as a predictor of success (Smolinski, Crawley, and Olsen 2017; Bochner et al. 2023; Chan et al. 2010). As such, we contend that an outbreak science approach to studying the components of response might be the first step towards developing the next generation of pandemic preparedness plans — with, ideally, a higher chance of success during the next pandemic.

## Data Availability

All data and code to reproduce our analysis are available on Github (github.com/cghss/epiintel).

https://github.com/cghss/epiintel

## ACKNOWLEDGEMENTS

We thank all collaborators in the development of the Georgetown Outbreak Activity Library, including Madison Alvarez, Aurelia Attal-Juncqua, Madison Berry, Matthew Boyce, Ellen Carlin, Stephanie Eaneff, Jessica Lin, Lauren McGivern, Jordan Schermerhorn, Emily Sherman, Claire Standley, Juliana St Goar, Hannah Todd, Kate Toole, Mark Wilcox, Aika Wojt, and Emily Woodrow. We thank our funders, who were not involved in the preparation or content of the study: Open Philanthropy supported the initial work on the Disease Outbreak News and the Georgetown Outbreak Activities Library (GOAL); PAX sapiens supported the analyses presented in this manuscript on epidemic intelligence. Finally, we thank all those who helped develop the GOAL resource, including participants in the collaborative workshops (David Brett-Major, Joseph Fair, Janice Hepburn, Jas Mantero, Ryan Morhard, and Rob Salerno) and those involved in data management and web interface design (Justin Kerr, Michael Van Maele, Kelsey Smith, and Nicole Stephan).

## References

Bochner, Aaron F., Issa Makumbi, Olaolu Aderinola, Aschalew Abayneh, Ralph Jetoh, Rahel L. Yemanaberhan, Jenom S. Danjuma, et al. 2023. “Implementation of the 7-1-7 Target for Detection, Notification, and Response to Public Health Threats in Five Countries: A Retrospective, Observational Study.” The Lancet. Global Health 11 (6): e871–79.

Carlson, Colin J., Matthew R. Boyce, Margaret Dunne, Ellie Graeden, Jessica Lin, Yasser Omar Abdellatif, Max A. Palys, Munir Pavez, Alexandra L. Phelan, and Rebecca Katz. 2023. “The World Health Organization’s Disease Outbreak News: A Retrospective Database.” PLoS Global Public Health, in press.

Chan, Emily H., Timothy F. Brewer, Lawrence C. Madoff, Marjorie P. Pollack, Amy L. Sonricker, Mikaela Keller, Clark C. Freifeld, Michael Blench, Abla Mawudeku, and John S. Brownstein. 2010. “Global Capacity for Emerging Infectious Disease Detection.” Proceedings of the National Academy of Sciences. https://doi.org/10.1073/pnas.1006219107.

Foy, Brian D., Kevin C. Kobylinski, Joy L. Chilson Foy, Bradley J. Blitvich, Amelia Travassos da Rosa, Andrew D. Haddow, Robert S. Lanciotti, and Robert B. Tesh. 2011. “Probable Non-Vector-Borne Transmission of Zika Virus, Colorado, USA.” Emerging Infectious Diseases 17 (5): 880–82.

Haldane, Victoria, Anne-Sophie Jung, Rachel Neill, Sudhvir Singh, Shishi Wu, Margaret Jamieson, Monica Verma, et al. 2021. “From Response to Transformation: How Countries Can Strengthen National Pandemic Preparedness and Response Systems.” BMJ 375 (November): e067507.

Katz, Rebecca, and Ellie Graeden. 2020. “Outbreak Activity Library: An Online, User-Friendly Compilation of Activities Essential for Effective Outbreak Response.” https://outbreaklibrary.org/export/Research%20Brief%202020-05%20-%20Essential%20activities%20for%20outbreak%20response.pdf.

Kreuder Johnson, Christine, Peta L. Hitchens, Tierra Smiley Evans, Tracey Goldstein, Kate Thomas, Andrew Clements, Damien O. Joly, et al. 2015. “Spillover and Pandemic Properties of Zoonotic Viruses with High Host Plasticity.” Scientific Reports 5 (October): 14830.

Laurenson-Schafer, Henry, Nikola Sklenovská, Ana Hoxha, Steven M. Kerr, Patricia Ndumbi, Julia Fitzner, Maria Almiron, et al. 2023. “Description of the First Global Outbreak of Mpox: An Analysis of Global Surveillance Data.” The Lancet. Global Health 11 (7): e1012–23.

Lloyd-Smith, James O., Dylan George, Kim M. Pepin, Virginia E. Pitzer, Juliet R. C. Pulliam, Andrew P. Dobson, Peter J. Hudson, and Bryan T. Grenfell. 2009. “Epidemic Dynamics at the Human-Animal Interface.” Science 326 (5958): 1362–67.

Phelan, Alexandra L., and Colin J. Carlson. 2022. “A Treaty to Break the Pandemic Cycle.” Science 377 (6605): 475–77.

Polonsky, Jonathan A., Amrish Baidjoe, Zhian N. Kamvar, Anne Cori, Kara Durski, W. John Edmunds, Rosalind M. Eggo, et al. 2019. “Outbreak Analytics: A Developing Data Science for Informing the Response to Emerging Pathogens.” Philosophical Transactions of the Royal Society of London. Series B, Biological Sciences 374 (1776): 20180276.

Rivers, Caitlin, Jean-Paul Chretien, Steven Riley, Julie A. Pavlin, Alexandra Woodward, David Brett-Major, Irina Maljkovic Berry, et al. 2019. “Using ‘outbreak Science’ to Strengthen the Use of Models during Epidemics.” Nature Communications 10 (1): 1–3.

Singh, Sudhvir, Christine McNab, Rose Mckeon Olson, Nellie Bristol, Cody Nolan, Elin Bergstrøm, Michael Bartos, et al. 2021. “How an Outbreak Became a Pandemic: A Chronological Analysis of Crucial Junctures and International Obligations in the Early Months of the COVID-19 Pandemic.” The Lancet 398 (10316): 2109–24.

Smolinski, Mark S., Adam W. Crawley, and Jennifer M. Olsen. 2017. “Finding Outbreaks Faster.” Health Security 15 (2): 215–20.

Wolfe, Nathan D., Claire Panosian Dunavan, and Jared Diamond. 2007. “Origins of Major Human Infectious Diseases.” Nature 447 (7142): 279–83.

